# Accurate diagnosis of apical hypertrophic cardiomyopathy using explainable advanced ECG analysis

**DOI:** 10.1101/2023.10.17.23297183

**Authors:** Rebecca K. Hughes, George D. Thornton, James W. Malcolmson, Iain Pierce, Shafik Khoury, Amanda Hornell, Kristopher Knott, Gabriella Captur, James C. Moon, Todd T. Schlegel, Martin Ugander

## Abstract

**Background:** Typical electrocardiogram (ECG) features of apical hypertrophic cardiomyopathy (ApHCM) include tall R waves and deep or giant T-wave inversion in the precordial leads, but these features are not always present. We tested whether explainable advanced ECG (A-ECG) could accurately diagnose ApHCM.

**Methods:** A-ECG analysis was performed on standard resting 12-lead ECGs in patients with ApHCM (n=75 overt, n=32 relative [<15mm hypertrophy]), a subgroup of which underwent cardiovascular magnetic resonance, n=92), and comparator subjects (n=2449), including healthy volunteers (n=1672), patients with coronary artery disease (n=372), left ventricular electrical remodelling (n=108), ischemic (n=114) or non-ischemic cardiomyopathy (n=57), and asymmetrical septal hypertrophy (ASH) HCM (n=126).

**Results:** Multivariable logistic regression identified four A-ECG measures that together discriminated ApHCM from other diseases with high accuracy (area under the receiver operating characteristics curve (AUC) [bootstrapped 95% confidence interval] 0.982 [0.965– 0.993]. Linear discriminant analysis also diagnosed ApHCM with high accuracy (AUC 0.989 [0.986–0.991]).

**Conclusion:** Explainable A-ECG has excellent diagnostic accuracy for ApHCM, even when the hypertrophy is relative, with A-ECG analysis providing incremental diagnostic value over imaging alone. The electrical (ECG) and anatomical (wall thickness) disease features do not completely align, suggesting future diagnostic and management strategies may incorporate both features.

## INTRODUCTION

The typical standard resting 12-lead electrocardiogram (ECG) in apical hypertrophic cardiomyopathy (ApHCM) has a distinctive appearance, with precordial tall R waves and deep T-wave inversion, with ‘giant’ negative T-waves (>10mm) in around half of cases^1^. However, whilst frequently characteristic, such ECG changes may vary over time, and electrically milder changes can be mistaken for other phenotypes or diseases (other HCM variants, other cardiomyopathies or myocardial ischemia)^2^. In initial disease descriptions from Japan, ApHCM diagnostic criteria were ECG-based with the use of vectorcardiography and with confirmation by invasive ventriculography^3^. Advances in imaging shifted this paradigm, and in current clinical practice, whilst the ECG may gatekeeper imaging diagnostic tests such as echocardiography and cardiovascular magnetic resonance (CMR), it is the imaging of apical hypertrophy that is the primary diagnostic criterion; a current de-emphasis of the electrical over anatomical phenotype components.

In left ventricular hypertrophy, the extent of secondary T-wave changes usually mirrors the amplitude of the QRS complex and R waves, and these have been shown to correlate in ApHCM^2,4^, with giant negative T waves and R-wave voltages >25mm having been demonstrated in those with more severe apical hypertrophy^5^. The typical ECG appearances are also seen in those with morphologically mild ApHCM – with <15mm apical hypertrophy but typical imaging features (loss of apical tapering, apical cavity obliteration in systole, ‘ace-of-spades’ appearance of left ventricular cavity, +/- apical microaneurysms), a subgroup termed relative ApHCM^6^. This not only calls into question the anatomical theory for the ECG changes, but also raises the question of whether the ECG changes that occur before the imaging phenotype fully develops could be relied upon for diagnosis.

The 12-lead ECG, standardized for almost 80 years^7^, was initially supplemented by labour intensive techniques such as vectorcardiography, but these have retreated from mainstream clinical use with the success of cardiac imaging. However, the emergence of electronic patient records with digital raw data storage of the ECG as standard, in combination with biobanks and advanced statistical methods, permit the return of reliable insights from ECGs and their delivery at healthcare system scale. Advanced ECG (A-ECG) uses derived three-dimensional vectorcardiograms^8^ and measures of QRS-wave and T-wave complexity^9^ to improve the diagnostic performance of the ECG in a number of domains, but not to date for ApHCM^10^. We sought to assess whether A-ECG analyses based on standard 12-lead ECG could accurately diagnose overt and relative ApHCM, and to assess relationships between electrical and anatomical changes.

## METHODS

The prospective ApHCM study was approved by the National Health Service Research Ethics Committee (NHS REC) and Health Research Authority (HRA) and conducted in accordance with the Declaration of Helsinki. All ApHCM subjects provided written, informed consent (REC 18/LO/0188 and 17/SC/0077). ECGs from other study participants recruited earlier from various centers around the world following informed consent or approval of a waiver of individual consent from a human subject ethics review board as previously described^10^.

### Study Populations

Patients with suspected or confirmed ApHCM/relative ApHCM (n=95) were prospectively recruited from tertiary referral cardiomyopathy clinics at St Bartholomew’s Hospital or St George’s University Hospital, London, UK, and underwent a standard resting 12-lead electrocardiogram where digital raw data was stored electronically, and a CMR scan. Three patients were later excluded due to the presence of a complete bundle branch block. A further 15 ApHCM subjects had the same ECG assessment but the imaging was echocardiography alone. Overt ApHCM was defined as maximum apical wall thickness (MWT) ≥15mm in end-diastole in conjunction with other characteristic features of the disease^11^ such as apical cavity obliteration, apical aneurysm, and suggestive ECG changes^4^. Relative ApHCM was defined previously^6^ as inappropriate apical hypertrophy compared to expected apical wall thickness but not exceeding 15mm, in combination with other characteristic features of the disease, as above. The diagnosis of overt disease used CMR or echocardiography, and relative ApHCM was diagnosed only by CMR.

Healthy subjects (n=1672) were defined as low risk, asymptomatic individuals with no cardiovascular or systemic disease, based on clinical history and physical examination. Exclusion criteria for healthy subjects included diagnosis of and treatment for hypertension or diabetes, current smoker, and increased blood pressure on examination (>140/90mmHg). They were recruited from the following centers: Johnson Space Center (USA), the Universidad de los Andes (Venezuela), the University of Ljubljana (Slovenia) and Lund University Hospital (Sweden)^10^.

Patients with established cardiovascular disease were subdivided into diagnostic groups: 1) occlusive coronary artery disease (n=372 – the presence of ≥50% obstructed in at least one major vessel by invasive coronary angiography, or, if angiography had not been performed, the presence of one or more reversible perfusion defects in a coronary artery territory on 99m-Tc-tetrofosmin single-photon emission computed tomography (SPECT), always with normal systolic function, 2) Left ventricular electrical remodelling (LVER, n=108), based on the presence of at least moderate LVH by imaging, but with normal systolic function, 3) ischemic and 4) non-ischemic cardiomyopathy (n=114, n=57), with left ventricular systolic dysfunction (ejection fraction ≤50%), or 5) asymmetrical septal hypertrophy (ASH) HCM (n=126). These groups were identified from the following centers: Texas Heart Institute (Houston, USA), the University of Texas Medical Branch (Galveston, USA), The University of Texas Health Science Center (San Antonio, USA), Brooke Army Medical Center (San Antonia, USA), St. Francis Hospital (Charleston, USA), the Universidad de los Andes (Mérida, Venezuela), and Lund Hospital (Lund, Sweden). ECGs were acquired within 30 days of the cardiac imaging examination.

### Electrocardiograms

For the 92/107 ApHCM subjects with contemporaneous CMRs, the resting 12-lead ECGs were performed on the day of CMR imaging using a Mortara ELI350 machine and stored in the patient’s electronic record (Cerner Millennium). These were extracted as DICOM files, pseudo-anonymized and converted to the xml file format for analysis. For the remaining 15 subjects, five-minute ECGs were performed. The other disease/control groups had a combination of 5-minute and 10-second ECGs. There were three analyses: Firstly, visual assessment of ECG DICOMs seeking the typical 12-lead ECG features of ApHCM – specifically tall R-waves (precordial leads [V1-V6] ≥14mm) and associated T-wave inversion (≥3mm). Secondly, conventional ECG measures of scalar durations, amplitudes included the following: 12-lead voltage, Cornell voltage (the sum of the S wave in V3 plus the R wave in lead aVL, where ECG LVH is defined as >2.8mV for males and >2.0mV for females^12^), Sokolow-Lyon criteria (the sum of the S wave in V1 plus the larger of the R wave in V5 or V6, where ECG LVH was defined as a >3.5mV^10,13^ Thirdly, advanced ECG analysis involved derived vectorcardiographic (VCG) and polarcardiographic measures of the planar and three-dimensional spatial angles, directions (azimuths and elevations) and magnitudes of the electrical activation pattern, and QRS- and T-wave complexity measures quantified by singular value decomposition (SVD).

### CMR acquisition and analysis

ApHCM subjects underwent CMR including mapping and late gadolinium enhancement (LGE) to exclude phenocopies. Scans were performed at Barts Heart Centre and Chenies Mews Imaging Centre on a 1.5 Tesla magnet (Aera, Siemens Healthcare, Erlangen, Germany) using standard clinical protocols. CMRs were analysed using commercially available software (CVI42, Circle Cardiovascular Imaging, Calgary, Canada). LV volume analyses used a validated machine learning algorithm^14^, as did MWT^15^. LGE was quantified using the full-width half-maximum (FWHM) technique with LGE expressed in grams and as a % of total myocardium. An apical aneurysm (≥5mm) or micro-aneurysm (<5mm) was defined by the presence of an akinetic/dyskinetic motion, scarring and a non-obliterating apical cavity typically distal to an area of obliteration.

### Statistical analysis

Statistical analysis was performed with SAS JMP 11.0 (Cary, NC) and R version 4.1.2 (R Foundation for Statistical Computing, Vienna, Austria) with packages MASS for linear discriminant analysis (LDA), and multiROC for receiver operator curve analysis. Normality was assessed visually on histograms and using Kolmogorov-Smirnov test. Normally distributed and not-normally distributed continuous data were presented as mean±standard deviation or median [interquartile range], respectively, and compared across participant groups using the independent Student’s *t*-test or Mann-Whitney-Wilcoxon test, as appropriate. Categorical data were presented as counts and percentage and compared using chi-square test. Correlation was assessed with Pearson’s or Spearman’s coefficient for normally and non-normally distributed data respectively. A p-value <0.05 was considered statistically significant. Multivariable logistic regression analysis was performed to see which A-ECG measures, when combined, differentiated ApHCM from all other patients, either diseased or healthy. Measures demonstrating significant differences by univariate analyses were then subjected to multivariable logistic regression analysis by using standard stepwise procedures. LDA was also performed to test the accuracy of distinguishing each disease, including ApHCM, from every other disease, and from health through similar A-ECG-related feature selection. To determine the diagnostic accuracy of both the logistic regression and the LDA, the areas under the receiver operating characteristics curve (AUC) were calculated, and bootstrap resampling was performed 3000 times to estimate prospective performance and to obtain 95% confidence intervals (CI) for sensitivity, specificity and AUC.

## RESULTS

107 subjects with confirmed overt or relative ApHCM had a 12-lead ECG available for A-ECG analysis. Of these, 92/107 had contemporaneous CMR. 60/92 (65%) were classified as overt ApHCM (apical MWT ≥15mm, age 58±13 years, BSA 1.92±0.20m^2^, 75% male) and 32/92 (35%) as relative ApHCM (apical MWT <15mm but other characteristic features of the disease, as described above, age 56±14 years, BSA 1.88±0.13m^2^, 78% male). The demographic, baseline ECG and CMR characteristics of the ApHCM subjects are detailed in **Table 1**.

**Table 1.**
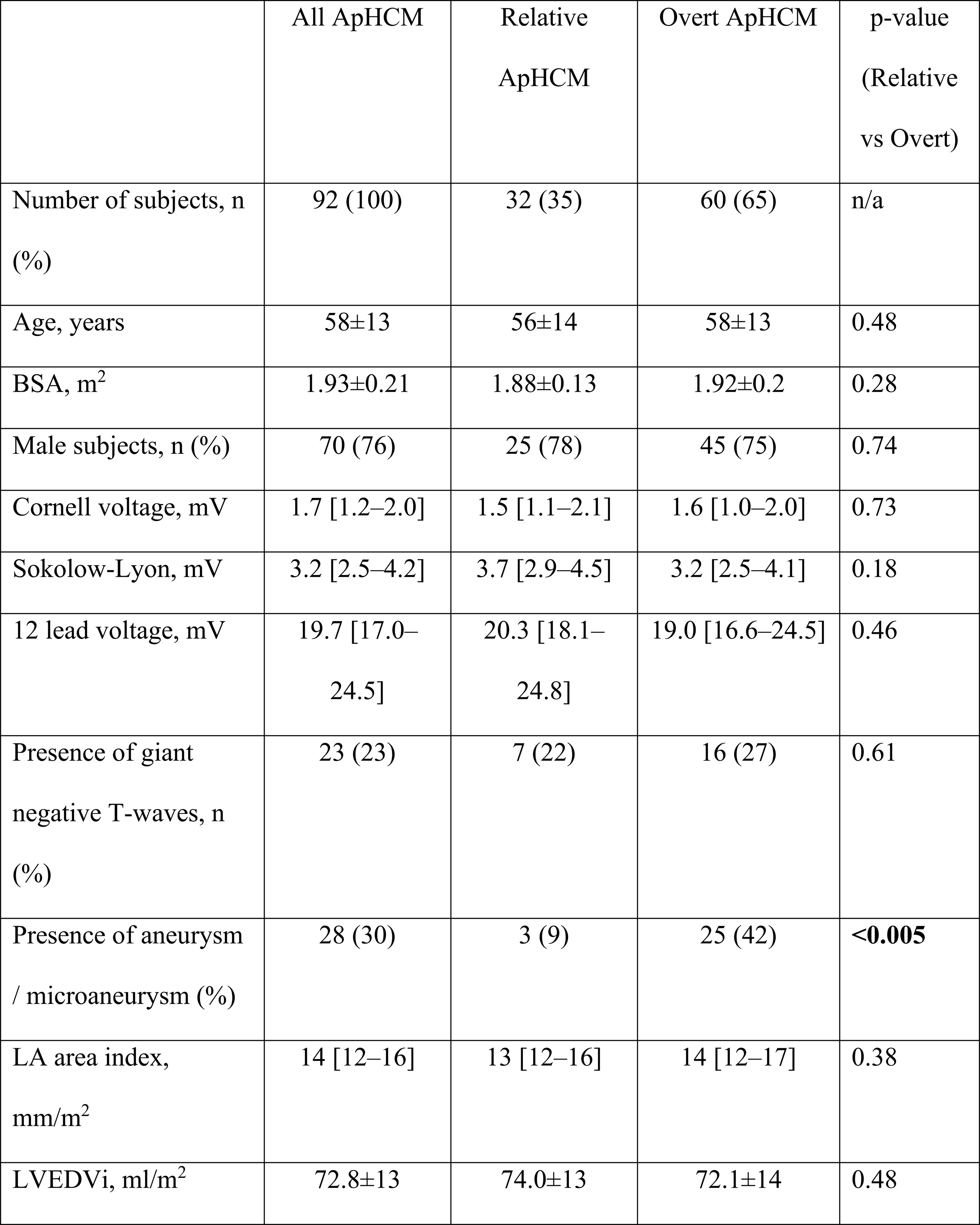
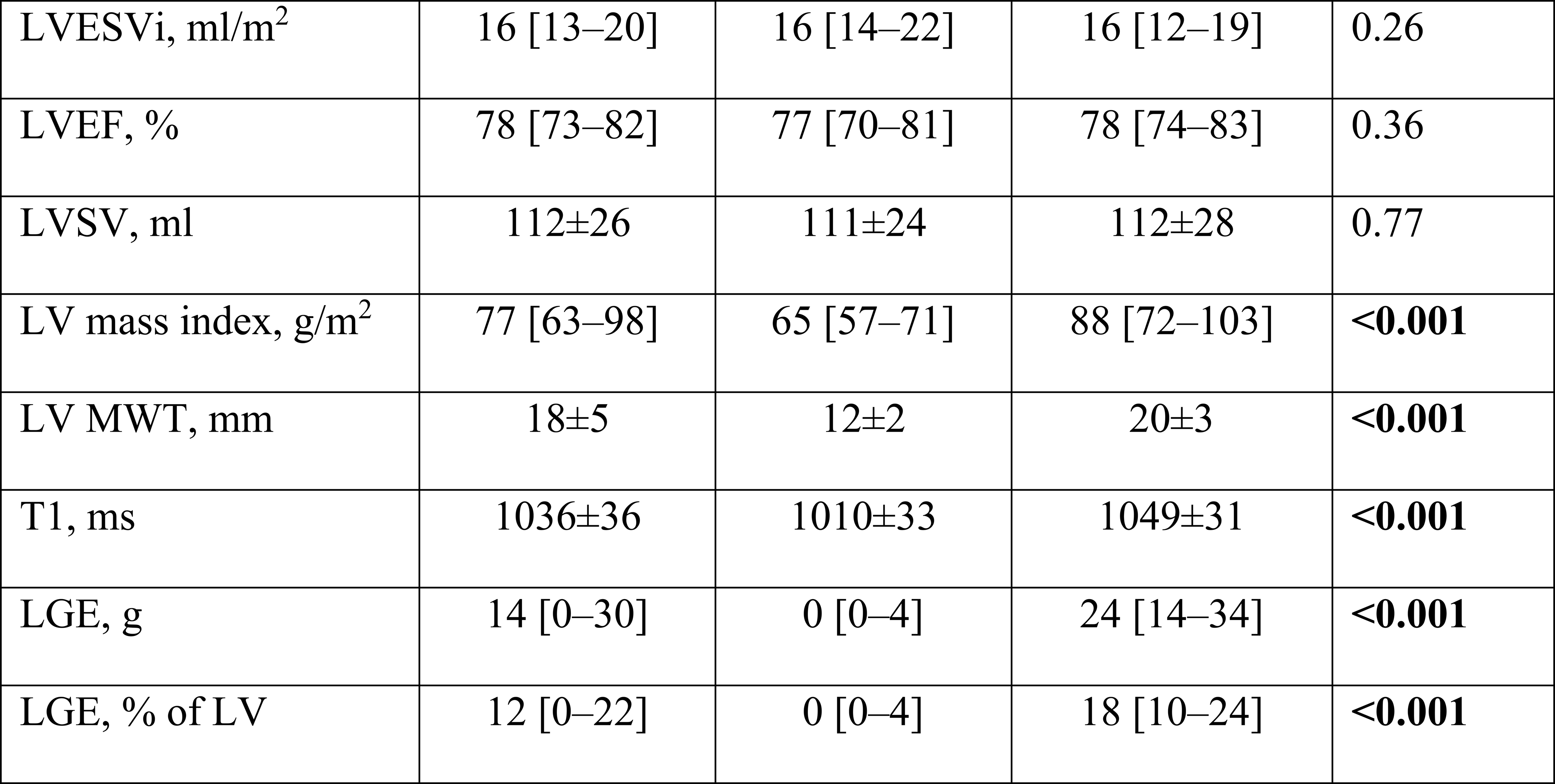
Comparison of baseline demographics, standard ECG characteristics, and CMR characteristics of relative vs overt ApHCM.

### Amplitude criteria

Visual ECG assessment was performed on the 92 ApHCM subjects with contemporaneous CMRs. 78/92 (85%) subjects had typical ECG features by amplitude criteria of ApHCM. Of the remaining 14 subjects with atypical ECG features, 5/14 had relative ApHCM and 9/14 overt disease by CMR. 4/14 had apical aneurysms/microaneurysms and 10/14 did not. 53/78 (68%) of those with typical ECG changes had apical scar with a total scar burden of 13 [0-22] % of the left ventricle. 9/14 (64%) of those with atypical ECG features had apical scar and a total scar burden of 9 [0-21] % of the left ventricle (p=0.76). **Figure 1** shows a comparison of ECGs in ApHCM patients with typical and atypical ECGs, with corresponding CMR images.

**Figure 1.**
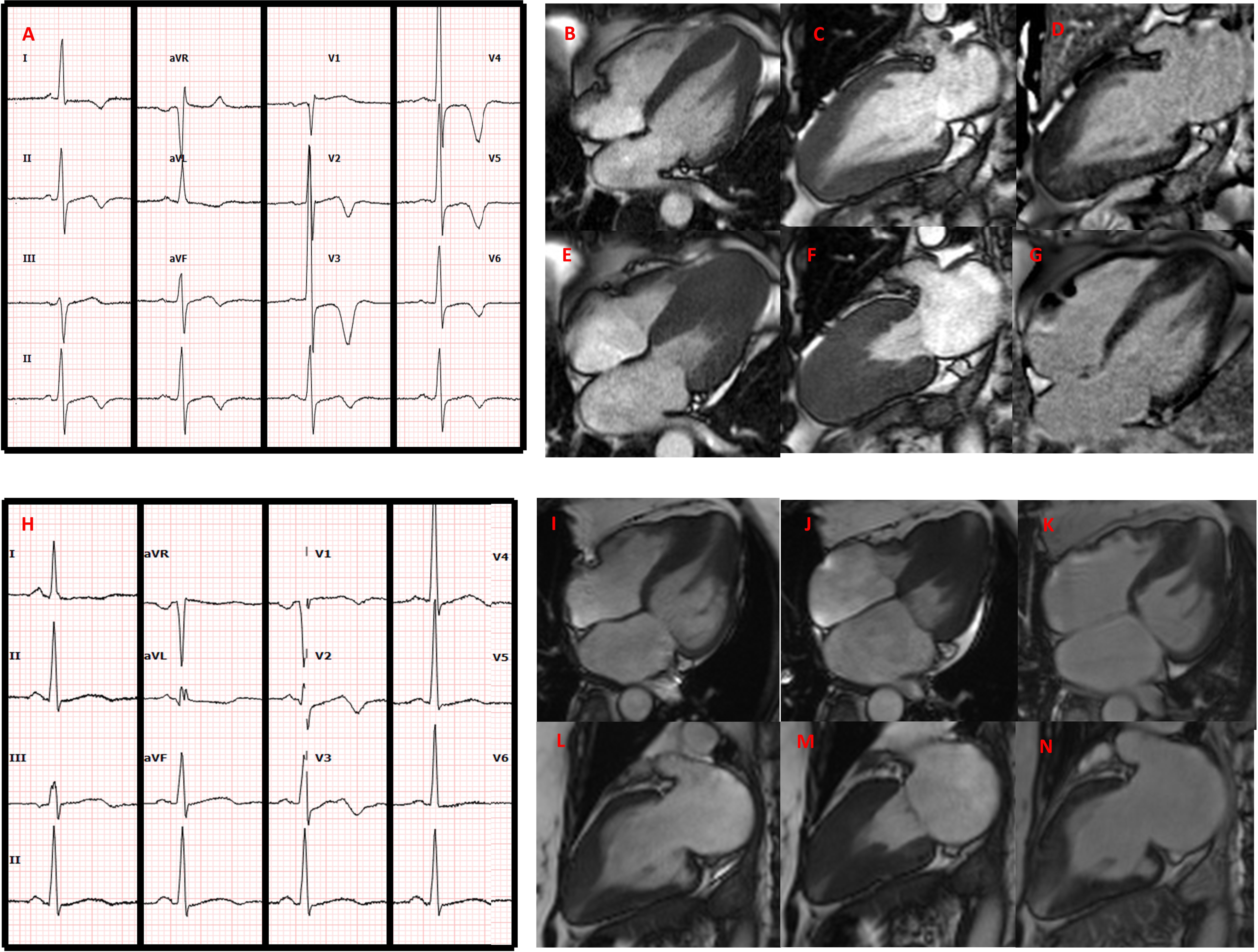
Typical vs atypical electrocardiogram (ECG) features in two patients with apical hypertrophic cardiomyopathy with corresponding cardiac magnetic resonance (CMR) imaging. ECG demonstrating visually typical ECG changes for ApHCM (A). Corresponding CMR demonstrating end-diastolic 4-chamber (B) and 2-chamber (C) views showing typical apical hypertrophy. End-systolic 4-chamber (E) and 2-chamber (F) views demonstrate significant apical cavity systolic obliteration. Patchy areas of scar are shown in late gadolinium enhancement (LGE) views (D, G). In contrast, atypical ApHCM ECG appearances are seen in H, with CMR demonstration of apical hypertrophy (I, J) in end-diastole, with the presence of an apical aneurysm in end-systole (L, M). Minimal apical scar seen on LGE views (K, N).

### A-ECG features

Multiple ECG variables differed between individuals with ApHCM versus those without in univariable analysis. However, multivariable analysis revealed that the following four ECG measures, when combined, had the best ability to distinguish ApHCM from all other pathologies and from health: (1) the direction of the peak of the T-wave loop in the vectorcardiographic horizontal plane; (2) the spatial peaks QRS-T angle; (3) the natural logarithm of the amplitude of the second eigenvector of the T wave after SVD; and (4) the depth of the Q wave in derived vectorcardiographic lead Z (**Table 2**). The AUC [bootstrapped 95% confidence interval] of the multivariable logistic regression score incorporating these four measures was 0.982 [0.965–0.993] (**Table 3**).

**Table 2.**
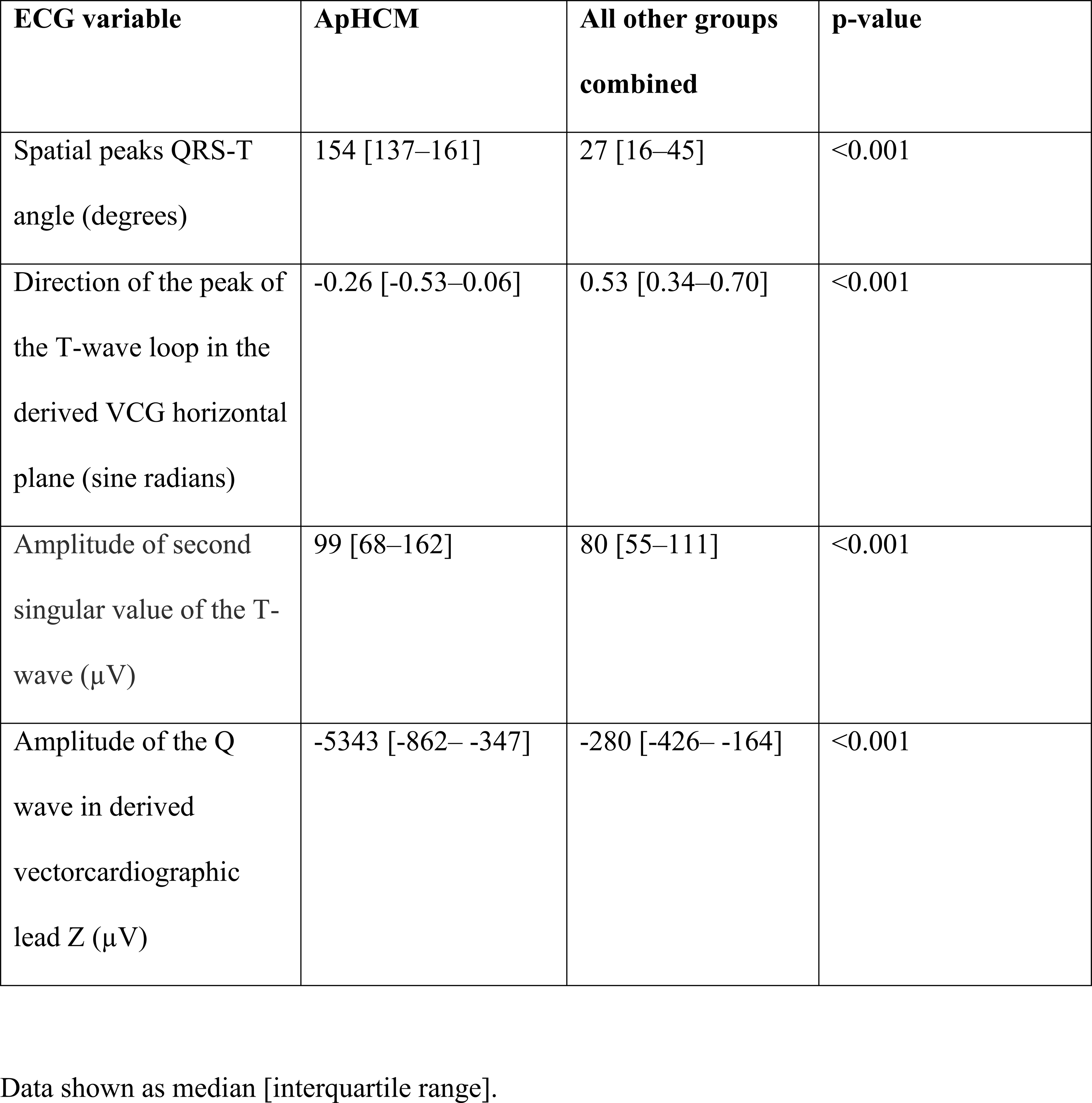
A-ECG measurements in ApHCM compared with all other healthy or diseased groups combined, including p-values from the logistic regression score for their discrimination.

**Table 3.**
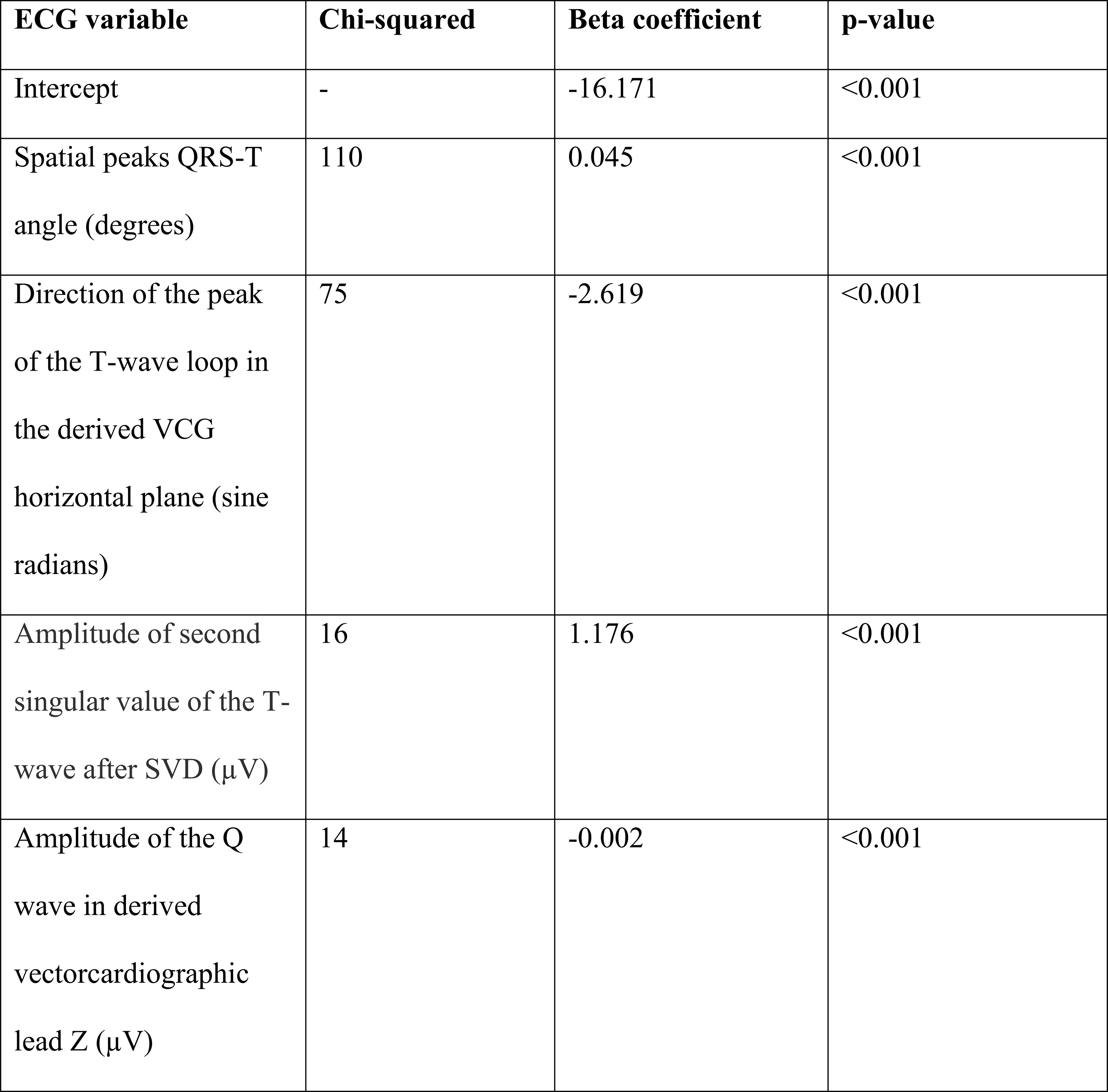
Strength of association (chi-squared) and beta coefficient for calculating a multivariable logistic regression score for optimally differentiating between ApHCM and all other diagnosis groups, whether healthy or diseased.

### LDA performance

LDA was also performed to determine not only the extent to which A-ECG could distinguish ApHCM from other disease conditions, but also the other conditions from one another. Thirty parameters, all with both univariate and final model-related individual p-values of <0.001, were included in a final LDA model, with overall accuracy of 91.6% (2342/2556), sensitivity of 88.8% and specificity of 99.3%, AUC of 0.989 [0.986-0.991], **Table 4**, **Figure 2**. Overall LDA performances for separating the other disease conditions from one and another and from cardiac health, are detailed in **Table 5**.

**Figure 2.**
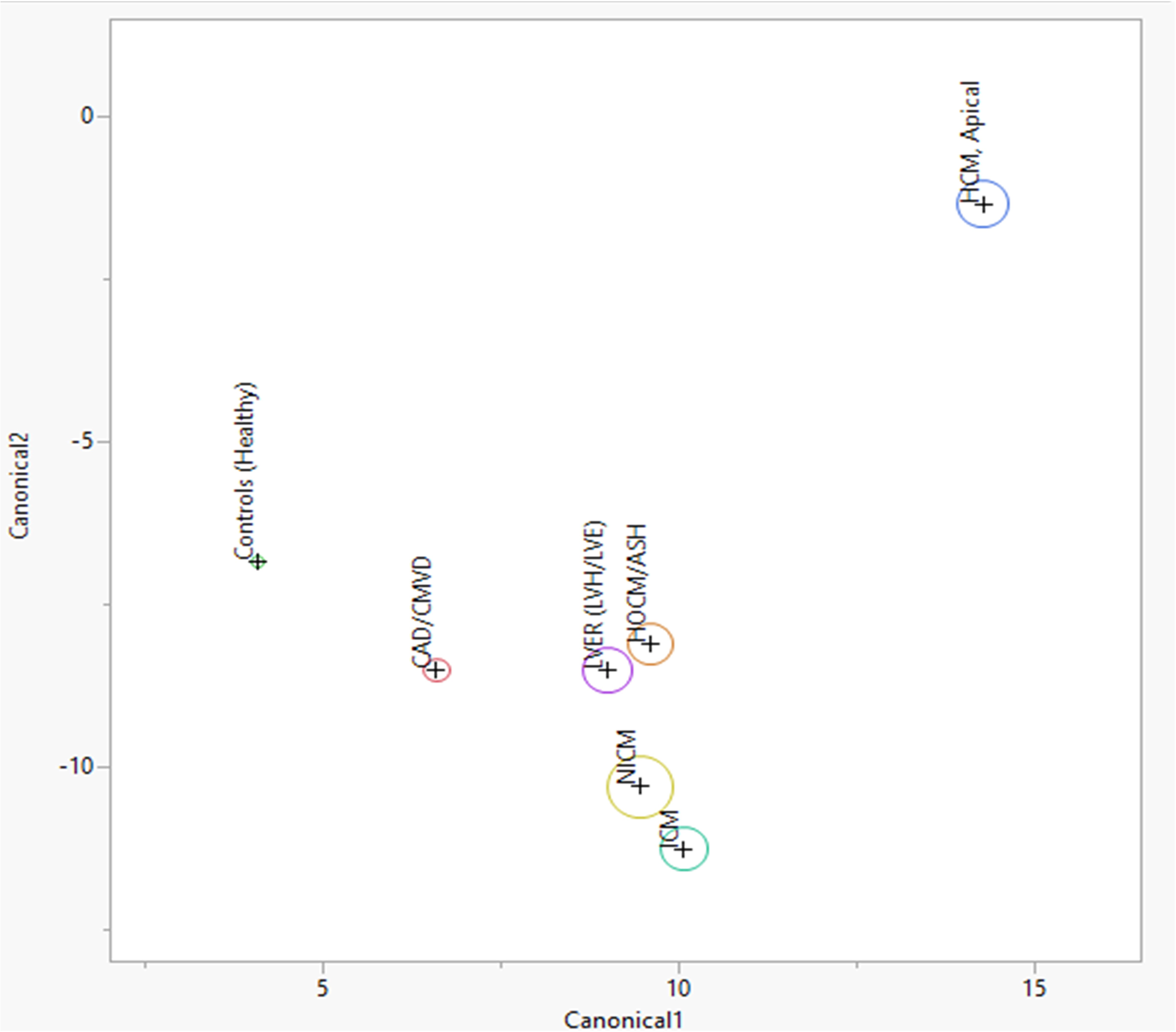
LDA projection of ApHCM diagnosis. Canonical plot demonstrating the clear distinction of ApHCM from the other disease groups and from health by LDA. The circle size represents the 95% confidence interval for the mean location of the given group. Note how this plot illustrates the distinctness of ECG characteristics for different diagnoses. CAD/CMVD = coronary artery disease/coronary microvascular disease; ICM = ischemic cardiomyopathy; HCM, apical = apical hypertrophic cardiomyopathy; HCM/ASH = asymmetrical septal hypertrophic cardiomyopathy; LVER (LVH/LVE) = left ventricular electrical remodelling; NICM = non-ischemic cardiomyopathy.

**Table 4.**
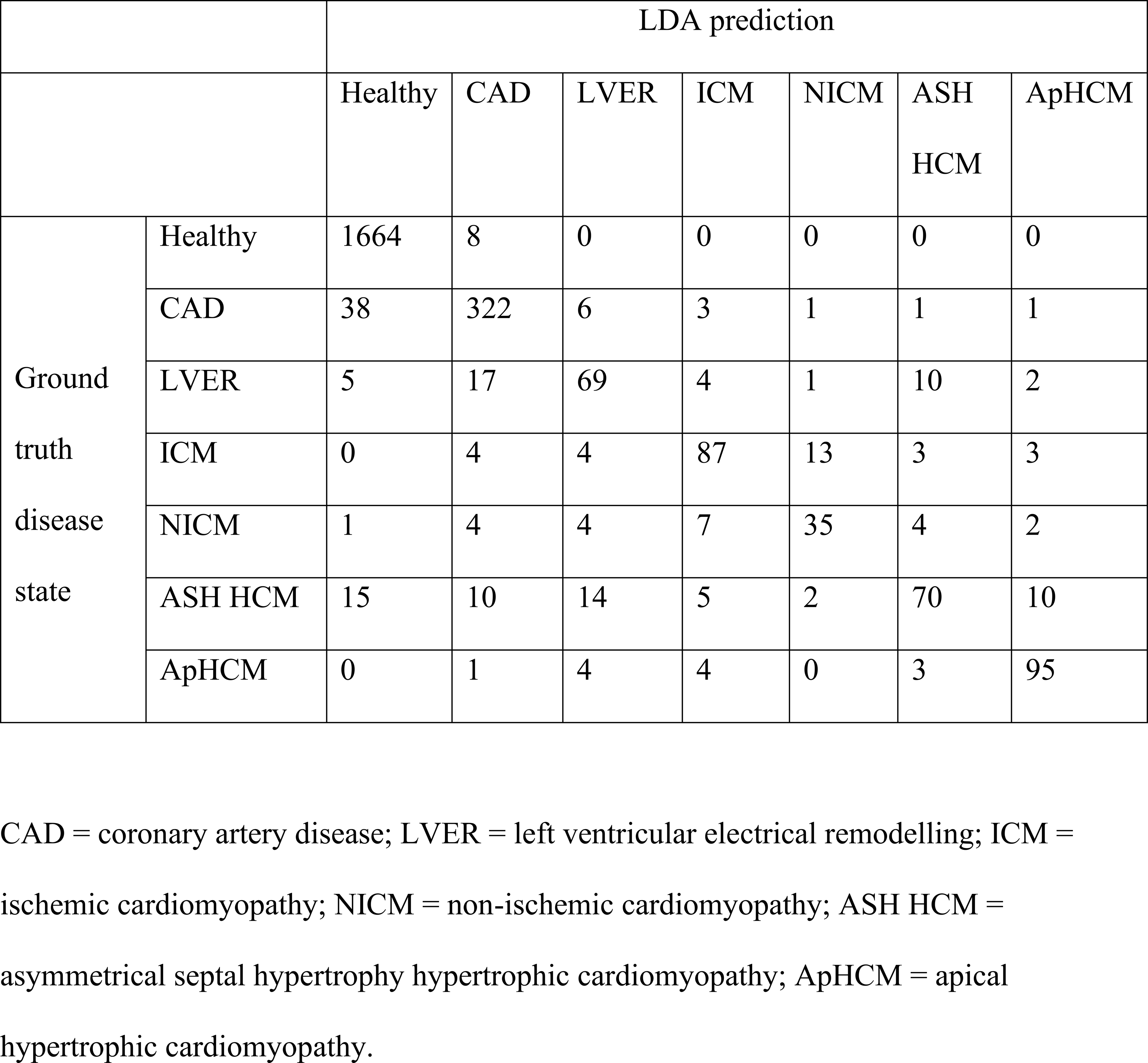
Diagnostic performance of linear discriminant analysis for predicting the respective ground truth diagnoses. Numbers denote numbers of individuals out of the full population (n=2451).

**Table 5.**
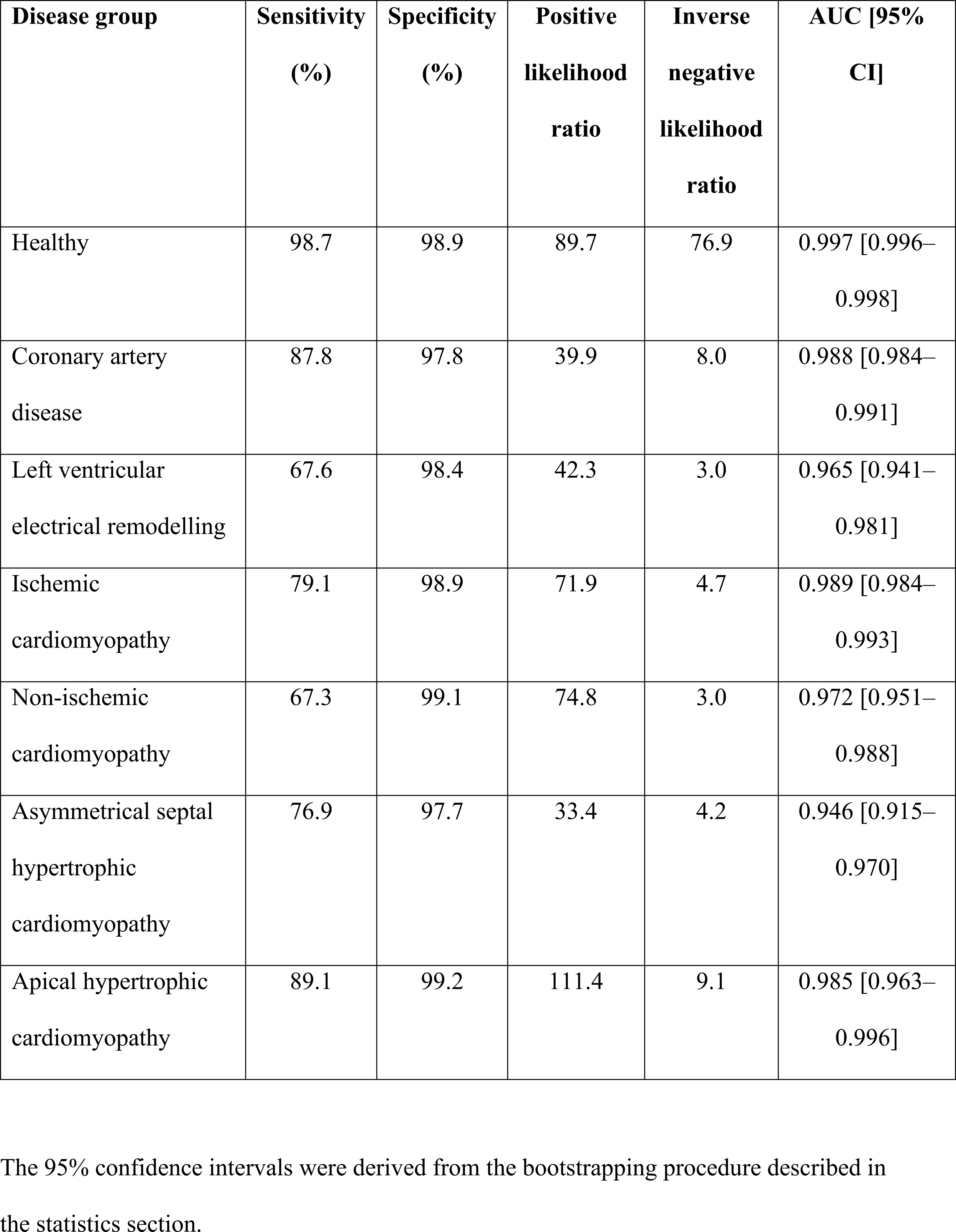
Linear discriminant analysis performance for identifying the respective diseases.

### LDA misclassification

12 subjects with ApHCM (6 relative, 6 overt) were misclassified as another diagnosis, namely: coronary artery disease in 1 subject with a large apical aneurysm, ischemic cardiomyopathy in 4 subjects (one with ApHCM and an apical microaneurysm, one with ApHCM and an apical aneurysm and two with relative ApHCM), left ventricular electrical remodelling in 4 subjects (three with relative ApHCM, one with ApHCM), and ASH HCM in three subjects (one with mixed septal and apical disease, one with ApHCM and apical aneurysm, one with relative ApHCM). No subjects with ApHCM were misclassified as healthy. 11/12 of these subjects had visually atypical resting 12-lead ECGs by our proposed amplitude criteria. The subject with typical visual ECG appearances (with a maximum R-wave amplitude of 25mm and maximum T-wave depth 7mm) had overt ApHCM with a MWT 22mm and significant apical LGE and was misclassified as ischemic cardiomyopathy. This patient had also had a contemporaneous coronary computerized tomography angiogram which showed only mild, non-obstructive coronary artery disease.

Of the remaining 2449 subjects, comprising the other healthy or disease groups studied in the LDA, 19 were misclassified as having ApHCM (0.8%). Of these, one had coronary artery disease, two had left ventricular electrical remodelling, three had ischemic cardiomyopathy, two had non-ischemic cardiomyopathy, and 11 had ASH HCM (all of whom had a degree of apical hypertrophy). No healthy volunteers were misclassified as having ApHCM.

### Vectorcardiographic features of ApHCM

Within both the logistic regression and the LDA, the direction of the peak of the T-wave loop in the derived vectorcardiographic horizontal (transverse) plane and the spatial peaks QRS-T angle were the most discriminative variables. In ApHCM, the QRS loop is typically directionally normal, but with increased QRS voltages, anecdotally suggested to manifest in increased Sokolow-Lyon voltage on the conventional ECG^16^. The main driver of increased spatial peaks QRS-T angle in ApHCM is therefore an abnormally directed T-wave loop, with abnormal rightward displacement of the T loop in the frontal and horizontal planes, and abnormal posterior displacement in the left sagittal plane.

By comparison, in the current study ASH HCM more often had abnormally directed QRS loops, especially excessively posteriorly-directed QRS loops in the horizontal plane, a reflection of pathological left ventricular electrical remodelling. This finding is often also accompanied by an initial rightward septal delay at 30ms into the QRS loop, and with Cornell QRS voltages typically being more abnormally high than Sokolow-Lyon QRS voltages. The T-wave loops in ASH HCM were usually less abnormally directed than in ApHCM, often being only modestly more anterior than normal in the horizontal plane, and rarely as overtly rightward as those in ApHCM (**Figure 3**). Furthermore, for purposes of comparison with previous ECG findings in ApHCM^2^, the R-wave amplitude in V5 was shown to correlate with T-wave depth in V5 in ApHCM patients in this study (r=-0.72, p<0.001).

**Figure 3.**
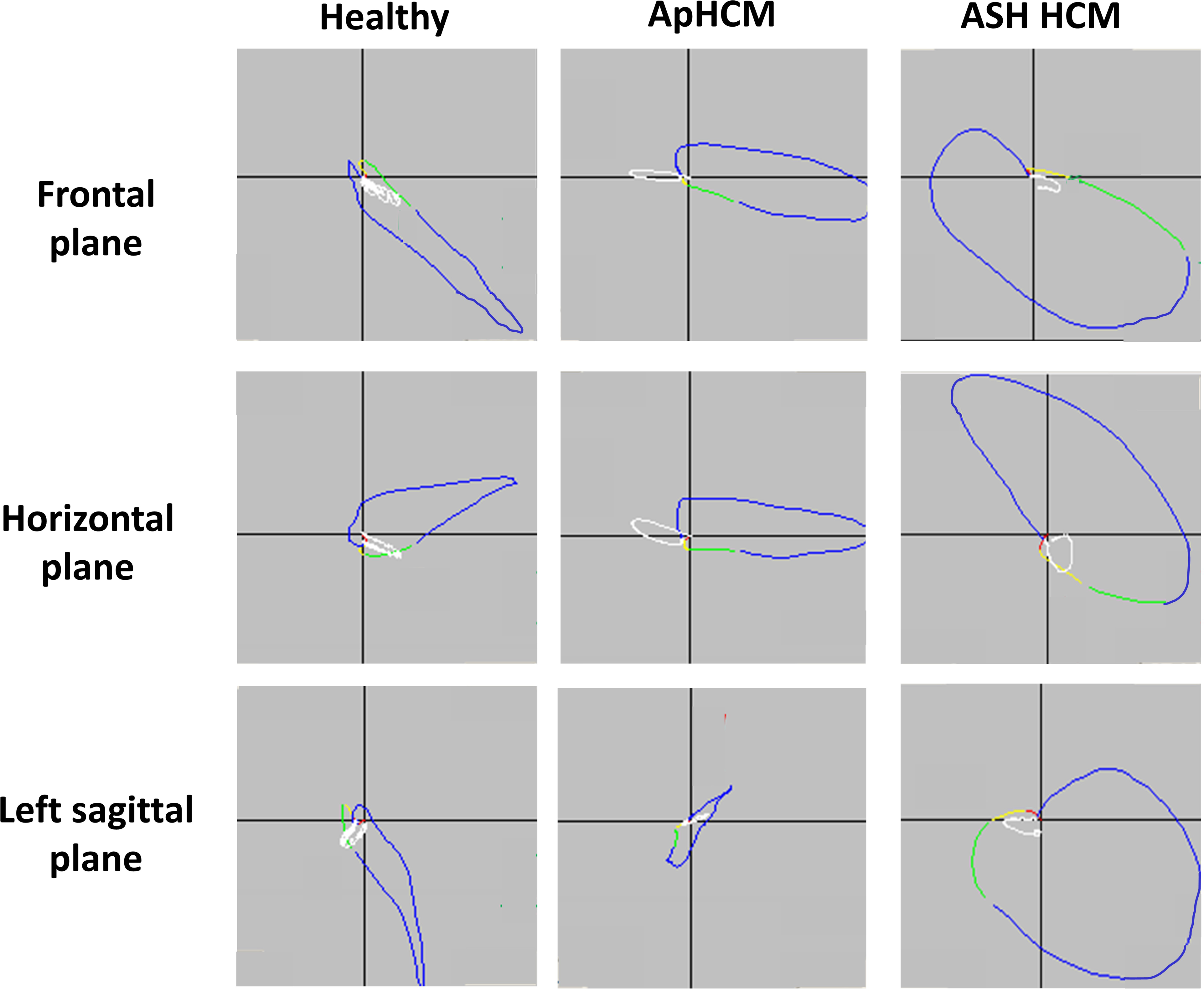
Vectorcardiographic appearances. Representative vectorcardiogram loops showing differences between the vectorcardiographic appearances of a single healthy subject (far left column) vs a patient with apical hypertrophic cardiomyopathy (ApHCM, middle column) and a patient with asymmetrical septal hypertrophy (ASH HCM, far right column). In healthy subjects, there is normal orientation of the QRS loops (in blue, with the initial direction of the loops shown in yellow and then green) and T-wave loops (in white). In ApHCM, there is typically a grossly abnormal rightward direction of the T-wave loops in the frontal and horizontal (transverse) planes and an abnormal posterior direction of the T-wave loop in the left sagittal plane. These severe T-wave directional changes result in notably increased spatial peaks QRS-T angles, even though QRS loops are mostly normally directed. By contrast, in ASH HCM compared to ApHCM, the T-wave loops are less abnormally directed in all planes, generally resulting in less abnormal spatial peaks QRS-T angles, although the T-wave loops are usually still directed abnormally anteriorly in the horizontal and left sagittal planes. By contrast, the QRS loops in ASH HCM are more abnormal than in ApHCM, often with the tell-tale excessive posterior deflection of the late portion of the QRS loop in the horizontal and left sagittal planes.

### Comparison of relative and overt ApHCM

To assess whether electrical differences exist between those who reach current anatomical diagnostic thresholds for ApHCM compared to those with relative ApHCM who do not, univariable analysis was used to investigate if any of the ECG variables within the optimal logistic regression equation predicted LV MWT (**Supplemental table 1**). None of the variables did.

### Apical aneurysms

28/92 (30%) ApHCM subjects had apical aneurysms or microaneurysms; with a similar proportion of those with relative (3/32; 9%) vs overt (9/60; 15%) ApHCM. Scar burden was greater in those with aneurysms vs without (23% [16-29] vs 6% [0-17], p=0.001). 4/28 (14%) had ECGs misclassified by the LDA vs 8/67 (12%) ApHCM subjects without aneurysms (p=0.75).

## DISCUSSION

The main finding of the study is that ApHCM can be reliably and accurately diagnosed by standard 12-lead ECG with A-ECG analysis. Although further cardiac imaging has a natural role in risk stratification, the standard ECG alone can be used to diagnose ApHCM, even in those with typical imaging features but less than the 15mm apical hypertrophy currently required for diagnosis (relative ApHCM). This is important, since currently those with suggestive ECG findings but <15mm apical hypertrophy are missing out on an important diagnosis and subsequent management due to not fulfilling imaging-based wall thickness criteria. However, with the presence of disease convincingly demonstrated by A-ECG, it rather suggests that this is an early or mild phenotype that should not be ignored. This argument is strengthened by the logistic regression analysis results, demonstrating that none of the four A-ECG measures that together best distinguished ApHCM from other diseases had an association with MWT. This highlights that electrical and anatomical features do not always align, and that the distinctive ECG features in ApHCM are not solely due to macroscopic apical hypertrophy.

As has been shown by others previously, we found a strong correlation between R-wave amplitude and T-wave depth. Tall R-waves in ApHCM were hypothesized to be due to the anatomical location of hypertrophy causing unopposed expression of depolarisation vectorial forces^4^. However, myocardial mass and diffuse myocardial fibrosis, quantifiable using CMR, have been shown to have independent and opposing effects upon ECG voltage measures of LVH^17^. Similarly, according to that view, T-wave changes would therefore be secondary, usually mirroring in extent the amplitude of QRS complexes and R waves, as in other forms of ventricular hypertrophy. Therefore it should be expected that large R waves are associated with large secondary T-wave changes with opposite vectorial orientations to the cardiac apex (superiorly, rightwards, and posteriorly)^4^. The strong correlation between R-wave amplitude and T-wave depth suggests that the two are indeed interlinked. However, given the poor association between the diagnostically most predictive A-ECG measures and wall thickness, there are likely other pathophysiological processes responsible for the ECG changes than previously thought. This is supported by findings that wall thickness alone could not explain T-wave inversion in those with ApHCM with septal involvement^18^. The authors hypothesized that ionic remodelling in ApHCM results in longer action potential duration, delayed repolarization, and inverted T waves, but found that ionic remodelling had no effect on the QRS complex in the same group. However, this explanation offers little to explain the tall R-waves.

The LDA performed with a high accuracy, and has further diagnostic uses beyond its ability to diagnose ApHCM. Specifically, it distinguished all forms of heart disease from one another, and from health, with very good diagnostic accuracy. Importantly, in those where the LDA misclassified ApHCM into another group, no patient was misclassified as being healthy. Typical ECG features defined as the proposed ‘amplitude criteria’ on visual read were present in 85% of subjects, and the LDA accurately identified ApHCM in 89% of subjects. Although these proportions are similar, A-ECG is still advantageous as visual ECG assessment is not diagnostic for ApHCM, and the proposed LDA results have demonstrated excellent diagnostic capabilities. It also offers a quantitative characterisation of the ECG findings in ApHCM relative to other pathologies.

## LIMITATIONS

The dataset was not divided into training and test sets, but instead cross-validated through bootstrap resampling. Although such resampling allows for estimation of prospective accuracy within reasonable confidence limits, fully prospective validation is ultimately required. Moreover, the diagnosis of ApHCM by A-ECG has also not yet been determined to confer any additional prognostic information, and such studies are justified. The number of ApHCM patients with apical aneurysms was small and further training of the model with more aneurysmal ApHCM data might be needed to avoid the misclassification that arose in this group.

## CONCLUSION

ApHCM is as much an electrical as an imaging phenotype and can be accurately diagnosed using a standard 12-lead ECG with A-ECG analysis even in those with typical ECG features but less than the current 15mm apical hypertrophy required for diagnosis. Electrical (ECG) and anatomical (wall thickness) findings do not necessarily align, and macroscopic apical hypertrophy alone is not wholly responsible for ECG changes and thus should not be relied upon in isolation for diagnosis and management.

## Data Availability

Data can be provided upon reasonable request.

## NON-STANDARD ABBREVIATIONS AND ACRONYMS

A-ECG: Advanced electrocardiogram
ApHCM: Apical hypertrophic cardiomyopathy
ASH: Asymmetrical septal hypertrophy
AUC: Area under the curve
CMR: Cardiac magnetic resonance
ECG: Electrocardiogram
FWHM: Full-width half-maximum
HCM: Hypertrophic cardiomyopathy
HRA: Health Research Authority
LDA: Linear discriminant analysis
LGE: Late gadolinium enhancement
LV: Left ventricle / ventricular
LVER: Left ventricular electrical remodelling
LVH: left ventricular hypertrophy
MWT: Maximum wall thickness
NHS REC: National Health Service Research Ethics Council
SPECT: Single-photon emission computerized tomography
SVD: Singular value decomposition
VCG: Vectorcardiogram

## ACKNOWLEDGEMENTS

Nil

## CLINICAL PERSPECTIVE

### What is new?

- This study looks at the diagnostic power of the ECG in apical hypertrophic cardiomyopathy and demonstrates that a 12-lead ECG with advanced ECG analysis can be used to reliably diagnose the disease.
- It demonstrates that ECG changes occur early in the disease, before the development of overt hypertrophy.
- The most distinguishing A-ECG features in apical hypertrophic cardiomyopathy are not associated with maximum wall thickness, implying that there is more than just hypertrophy responsible for the characteristic ECG changes.

### What are the clinical implications?

- This allows clinicians to consider the diagnosis of apical hypertrophic cardiomyopathy if the patient has characteristic ECG changes, even if wall thickness criteria for HCM are not met.
- Reminds cardiologists of the diagnostic power of a simple 12-lead ECG and to consider revisiting vectorcardiography to gain greater electrical insights.

